# Role of Professionalism in the Policy Response to COVID-19: Does a Public Health or Medical Background Help?

**DOI:** 10.1101/2020.10.16.20213934

**Authors:** X. Li, W. Lai, Q. Wan, X. Chen

## Abstract

Less than 5 percent of Chinese cities had top-ranked officials with public health or medical backgrounds (PHMBGs). Does professionalism improve their response to a public crisis like the COVID-19 pandemic? Collecting résumés of government and Party officials in almost all prefectural Chinese cities, and matching with other data sources, including weather conditions, city characteristics, COVID-19-related policies, and health outcomes, we demonstrate that cities whose top officials had PHMBGs witnessed significantly lower infection rates, and often lower death rates, than cities whose top officials lacked such backgrounds. Mechanism testing suggests that the effects were at least partially explained by more rapid lockdown or community closure. Our findings offer insights into better preparation for future epidemics via improving leadership team composition, particularly recruiting major officials with PHMBGs.

**One Sentence Summary:** Cities whose top officials had PHMBGs saw lower infection and death rates, attributable to more rapid decision to lock down and close communities.

---

Coronavirus disease 2019 (COVID-19) was first reported in the city of Wuhan in Hubei province, China, in early December 2019. It then spread rapidly within China and worldwide, becoming a pandemic and bringing about devastating consequences (*1*). To contain the transmission of the novel coronavirus, countries around the world have adopted a wide range of stringent measures to reduce human interaction, such as lockdowns, closed-off community management, and large-scale quarantines and isolations.

The exponential growth of the virus highlights the importance of a swift response. However, COVID-19 countermeasures vary substantially in the timing of their implementation and their efficacy. For instance, most East Asian countries, with their existing contact-tracing systems and experience of viral outbreaks, could contain COVID-19 in a relatively short time. Western governments, on the other hand, often began with loose regulations and delayed responses, which were followed by blanket lockdowns. Countries that responded more quickly, such as Italy, Germany, Austria, and Switzerland, contained the spread of the virus more effectively than countries with a delayed response, including the United Kingdom, Sweden, and the United States (*2,3*). The effectiveness of the pandemic response has varied across countries and regions. For instance, the case fatality rate (CFR) in Italy at the time of writing is 13.98%, compared with only 5.40% in China (*4*). Response effectiveness also varies within countries. In China, Hubei province has a CFR of 6.62%, which is much higher than the average value of 0.82% for other Chinese provinces (*5*). In the early stage of COVID-19 infection, quarantine is often the most effective approach, especially in combination with other prevention and control measures (*6*). China’s early implementation of city lockdowns and closed-up community management may have avoided a large number of infections and casualties (*7*).

There is no doubt that combating the pandemic demands collaboration from the public, scientists, and bureaucrats. The characteristics of bureaucrats may be particularly important, as they oversee policy formulation and enforcement to contain the spread of the virus. Their scientific knowledge may affect their perceptions of the underlying risks and corresponding responses. Numerous studies have suggested that a more diverse leadership team tends to be more productive and creative in problem solving, and can lead to higher-quality decisions being made (*8,9*). Moreover, leaders’ educational backgrounds may shape their ideas and beliefs about policymaking (*10*). Leaders’ previous experience may also be a good predictor of their behavior (*11*). The ongoing COVID-19 pandemic offers the opportunity to better understand the role of officials’ professionalism in emergency response in a high-stakes context.

Do more competent officials strengthen the efforts to fight COVID-19? To test this hypothesis, we collected data on officials in 294 prefectural Chinese cities. We examined the effect of the officials’ professionalism, as measured by their having a public health or medical background (PHMBG), on epidemic control and prevention. A PHMBG was broadly defined as having received an education or worked in public health or medicine. Bureaucrats are central to social management and development in China. Professional leaders with a PHMBG may leverage their unique experience during a public health crisis to demonstrate urgently needed skills in monitoring infectious diseases and organizing an effective policy response (*12*). However, during the last four decades of China’s opening up and economic reform, economic growth has often been prioritized over public health, and only a small proportion of Chinese bureaucrats have a PHMBG. We collected the official résumés of Party secretaries and mayors, the officials responsible for administrating prefectural Chinese cities; from which we extracted information on their experiences relevant to a PHMBG. We merged this information with data from other sources, including COVID-19 infection and fatality data, a rich set of socioeconomic characteristics, and meteorological and environmental information at the prefectural city level that may influence virus transmission. Both local Party leadership (Party secretaries and deputy secretaries) and government leadership (mayor and deputy mayors) were included in the analysis (*13*). We estimated the effects of officials’ having a PHMBG on the COVID-19 infection rates, death rates, and public health measures taken. Our linear regression model at the city level controlled for covariates associated with both the officials’ profiles and virus transmission, including population density, GDP per capita, and number of doctors per 10,000 population. We also included weather controls, such as the average daily maximum temperature, average wind speed, and average precipitation, and the average air quality index (AQI).

Our results indicate that a Party secretary having a PHMBG was significantly associated with a lower infection rate for that official’s city. In addition, the backgrounds of the Party secretaries seemed to play a much bigger role than that of the mayors. We also found that having a PHMBG was pivotal for leaders in terms of the timely implementation of local public health measures, such as city lockdowns and community closure, to curb human-to-human virus transmission.

We also shed light on some important socioeconomic, environmental, and other factors that may influence virus transmission and policy response to COVID-19. We provide consistent evidence that a higher GDP per capita, as a measure of economic activity, was associated with a higher infection rate, while population density played little role (*14*). Our results also indicate that neither GDP per capita, nor air pollution exposure or population density was associated with mortality rates due to COVID-19 (*15-16*). We offer some evidence that female mayors were associated with a lower infection rate than their male counterparts. A lower educational attainment among both mayors and Party secretaries seemed to be associated with higher infection rates. Also, a lower educational attainment among mayors further increased the death rate due to COVID-19.

This analysis represents, to the best of our knowledge, the first attempt to empirically test the impact of officials’ professionalism on their policy response and performance in the face of the COVID-19 pandemic. We contribute to the literature in the following respects. First, we offer novel evidence of how officials’ backgrounds influence their competence in leading efforts to respond to a major public health crisis. Existing studies have mostly focused on the roles of officials’ gender, age, and educational attainment (*10, 17-19*). Second, we add to the literature on the determinants of virus transmission. In addition to socioeconomic factors, environmental conditions, the political system, and compliance with disease control norms (*16, 20*), the PHMBG of a political team has an important effect on transmission. However, this factor has been neglected in the literature. Third, examining China’s experience offers guidance for other governments on better handling the ongoing health crisis by improving the qualifications of major officials, especially in countries still in the midst of the pandemic.

This paper is organized as follows. The next section introduces the data and model used. The section entitled “Empirical Strategy” describes the regression analysis. The results section details the findings of the analysis. The final section presents the concluding remarks and recommends future possible research directions in this field.

## Data

We gathered data from several sources for this study. We manually collected the résumés of city officials, including secretaries, deputy secretaries, mayors, and deputy mayors, from the websites of all prefectural governments in China. These résumés provided information on age, gender, length of Party membership, education background, and work experience. We defined an official as having a PHMBG if they had received relevant education (e.g., they had studied medicine or public health) and/or worked in relevant institutions (e.g., hospital, public health department, center of disease control). Data on confirmed COVID-19 cases at the prefecture city level were obtained from Johns Hopkins CSSE Dashboard up to February 29, 2020 (*21*). Our main reason for focusing on epidemic control and prevention prior to March 2020 was that the COVID-19 pandemic had been brought largely under control in China (with the exception of Wuhan) by the end of February (*22*). We dropped Wuhan from our analysis because it was the place of origin and epicenter of the pandemic, so the quality of disease surveillance data and public health measures in that city may not have been representative. Some small cities, primarily distributed in Tibet, Xinjiang, Inner Mongolia, and Hainan, were also excluded due to a lack of socioeconomic information. Hong Kong, Macau, and Taiwan were not included because their administrative structures are different from those of mainland China, so they are not very comparable. Our final sample included 294 prefectural cities in China. We obtained socioeconomic and health data from the China City Statistical Yearbook 2019, including GDP per capita, population density, and number of doctors per 10,000 population. Each city’s average daily AQI of 2019 was obtained from the website of China National Municipal Ecological and Environmental Monitoring Center. We used historical AQI data to capture people’s respiratory health conditions. Daily meteorological data for each city (from January 19, 2020 to February 29, 2020), covering maximum temperature, precipitation, and wind speed, were collected from the U.S. Weather National Oceanic Atmospheric Administration. Weather variables are influential for virus spread. They are averaged to generate cross-sectional weather controls. We also collected data on the timing of city lockdowns and community closure based on official announcements and media reports.

Table 1 summarizes the data on the cities with officials without (left panel) and with (right panel) PHMBGs. The average confirmed cases and deaths per million population for cities whose officials had no PHMBG were 30.8 and 0.78, respectively, which are significantly higher than the figures for cities whose officials did have a PHMBG (24.3 and 0.33). Table 1 also displays salient variations in population density, GDP per capita, the AQI, average daily maximum temperature, and number of doctors across the left and right panels (i.e., with and without a PHMBG, respectively). For example, the number of doctors per 10,000 people ranged from 0.14 to 10.94 in the left panel, and from 0.02 to 7.64 in the right panel, indicating a large heterogeneity in healthcare capacity across cities in China. Table 1 also shows there was a much smaller share of females occupying the positions of Party secretaries or mayors than their male counterparts. The average age of the secretaries was around 55 years old; on average, they were 3 years older than the mayors. Cities whose officials had PHMBGs adopted lockdown or community closure earlier. The speed of implementation of these measures was measured by the number of days between January 23, 2020, i.e., the date when Wuhan initiated lockdown, and the date of implementation.

**Table 1.**
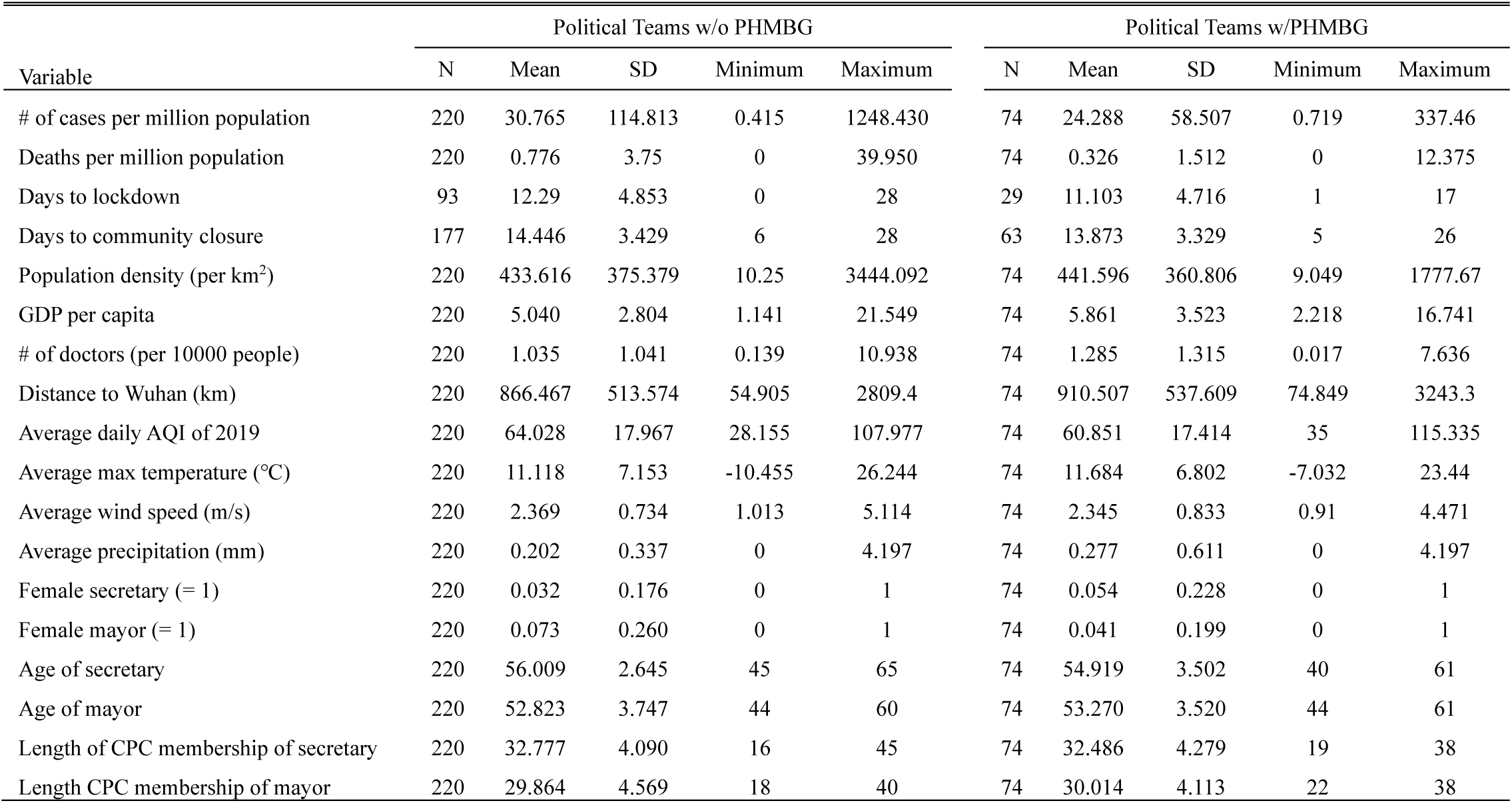
Summary Statistics (*32*).

| Variable | Political Teams w/o PHMBG |  |  |  |  | Political Teams w/PHMBG |  |  |  |  |
| --- | --- | --- | --- | --- | --- | --- | --- | --- | --- | --- |
|  | N | Mean | SD | Minimum | Maximum | N | Mean | SD | Minimum | Maximum |
| # of cases per million population | 220 | 30.765 | 114.813 | 0.415 | 1248.430 | 74 | 24.288 | 58.507 | 0.719 | 337.46 |
| Deaths per million population | 220 | 0.776 | 3.75 | 0 | 39.950 | 74 | 0.326 | 1.512 | 0 | 12.375 |
| Days to lockdown | 93 | 12.29 | 4.853 | 0 | 28 | 29 | 11.103 | 4.716 | 1 | 17 |
| Days to community closure | 177 | 14.446 | 3.429 | 6 | 28 | 63 | 13.873 | 3.329 | 5 | 26 |
| Population density (per km<sup>2</sup>) | 220 | 433.616 | 375.379 | 10.25 | 3444.092 | 74 | 441.596 | 360.806 | 9.049 | 1777.67 |
| GDP per capita | 220 | 5.040 | 2.804 | 1.141 | 21.549 | 74 | 5.861 | 3.523 | 2.218 | 16.741 |
| # of doctors (per 10000 people) | 220 | 1.035 | 1.041 | 0.139 | 10.938 | 74 | 1.285 | 1.315 | 0.017 | 7.636 |
| Distance to Wuhan (km) | 220 | 866.467 | 513.574 | 54.905 | 2809.4 | 74 | 910.507 | 537.609 | 74.849 | 3243.3 |
| Average daily AQI of 2019 | 220 | 64.028 | 17.967 | 28.155 | 107.977 | 74 | 60.851 | 17.414 | 35 | 115.335 |
| Average max temperature (°C) | 220 | 11.118 | 7.153 | -10.455 | 26.244 | 74 | 11.684 | 6.802 | -7.032 | 23.44 |
| Average wind speed (m/s) | 220 | 2.369 | 0.734 | 1.013 | 5.114 | 74 | 2.345 | 0.833 | 0.91 | 4.471 |
| Average precipitation (mm) | 220 | 0.202 | 0.337 | 0 | 4.197 | 74 | 0.277 | 0.611 | 0 | 4.197 |
| Female secretary (= 1) | 220 | 0.032 | 0.176 | 0 | 1 | 74 | 0.054 | 0.228 | 0 | 1 |
| Female mayor (= 1) | 220 | 0.073 | 0.260 | 0 | 1 | 74 | 0.041 | 0.199 | 0 | 1 |
| Age of secretary | 220 | 56.009 | 2.645 | 45 | 65 | 74 | 54.919 | 3.502 | 40 | 61 |
| Age of mayor | 220 | 52.823 | 3.747 | 44 | 60 | 74 | 53.270 | 3.520 | 44 | 61 |
| Length of CPC membership of secretary | 220 | 32.777 | 4.090 | 16 | 45 | 74 | 32.486 | 4.279 | 19 | 38 |
| Length CPC membership of mayor | 220 | 29.864 | 4.569 | 18 | 40 | 74 | 30.014 | 4.113 | 22 | 38 |

Figure 1 visualizes the spatial distribution of the prefectural city officials with PHMBGs. We classified the sampled cities into four categories: both the principal and deputy officials have a PHMBG; only the principal officials have a PHMBG; only the deputy officials have a PHMBG; and no city officials have a PHMBG. The figure shows no clear pattern of spatial clustering, indicating that city officials with a PHMBG may be quasi-randomly distributed. Figures 2a and 2b display the performance of epidemic control and prevention (confirmed cases per million population and deaths per million population), respectively, and Figures 2c and 2d show the timing of actions taken (city lockdown and community closure, respectively) in each city. Both Figure 1 and Figure 2 show large spatial variations across prefectural cities. To test the hypothesis that having a PHMBG is associated with officials enacting more timely policy response, as well as better epidemic control performance, we next report the results of our regression analysis and placebo tests.

**Fig. 1.**
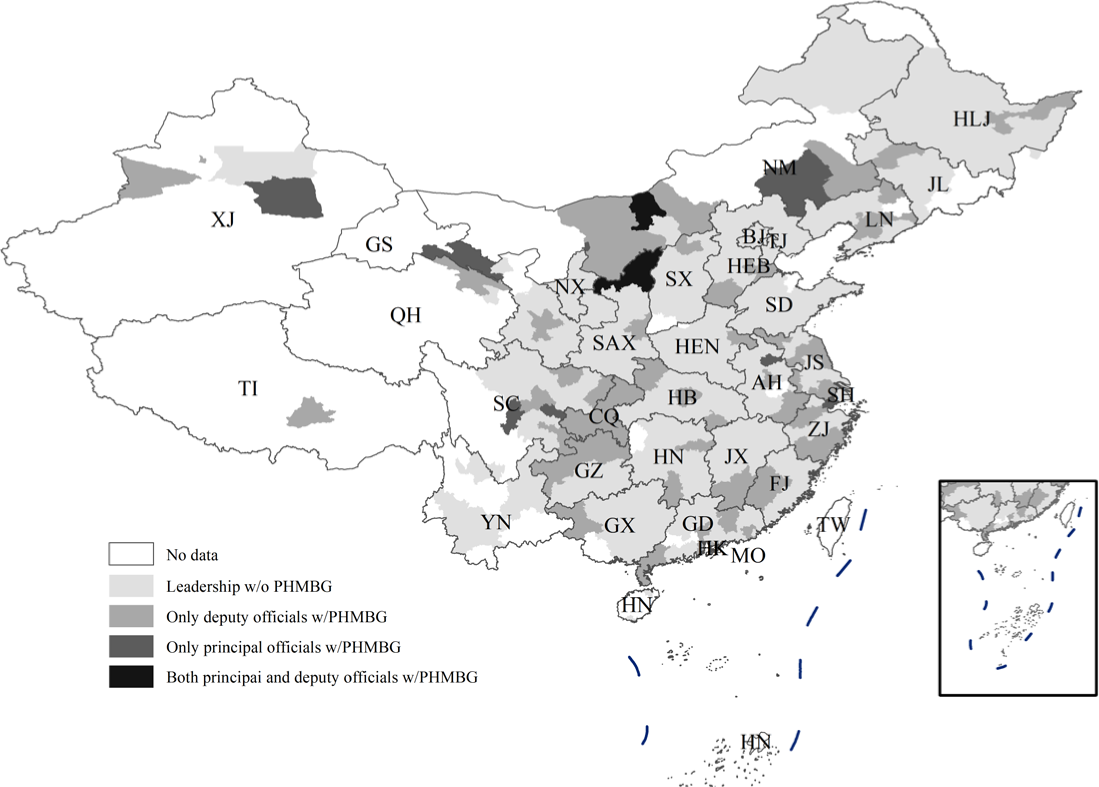
Public Health or Medical Background among Prefectural City Officials (*28*).

**Fig. 2.**
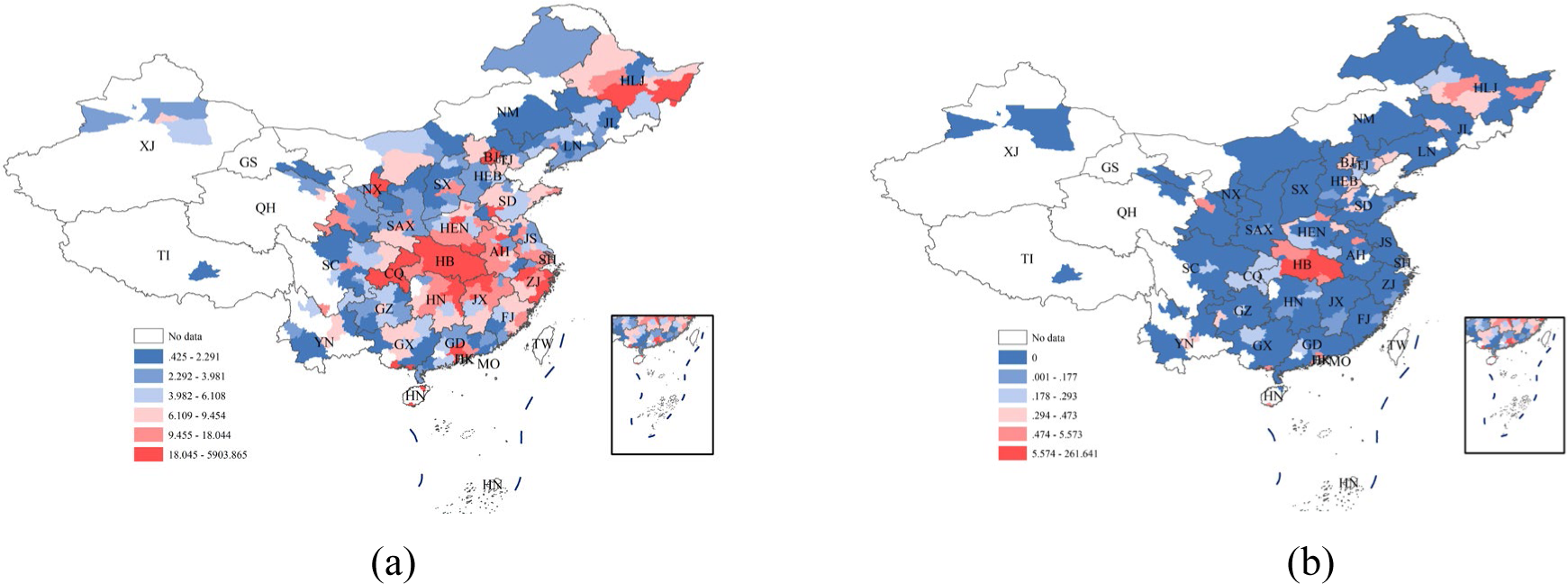
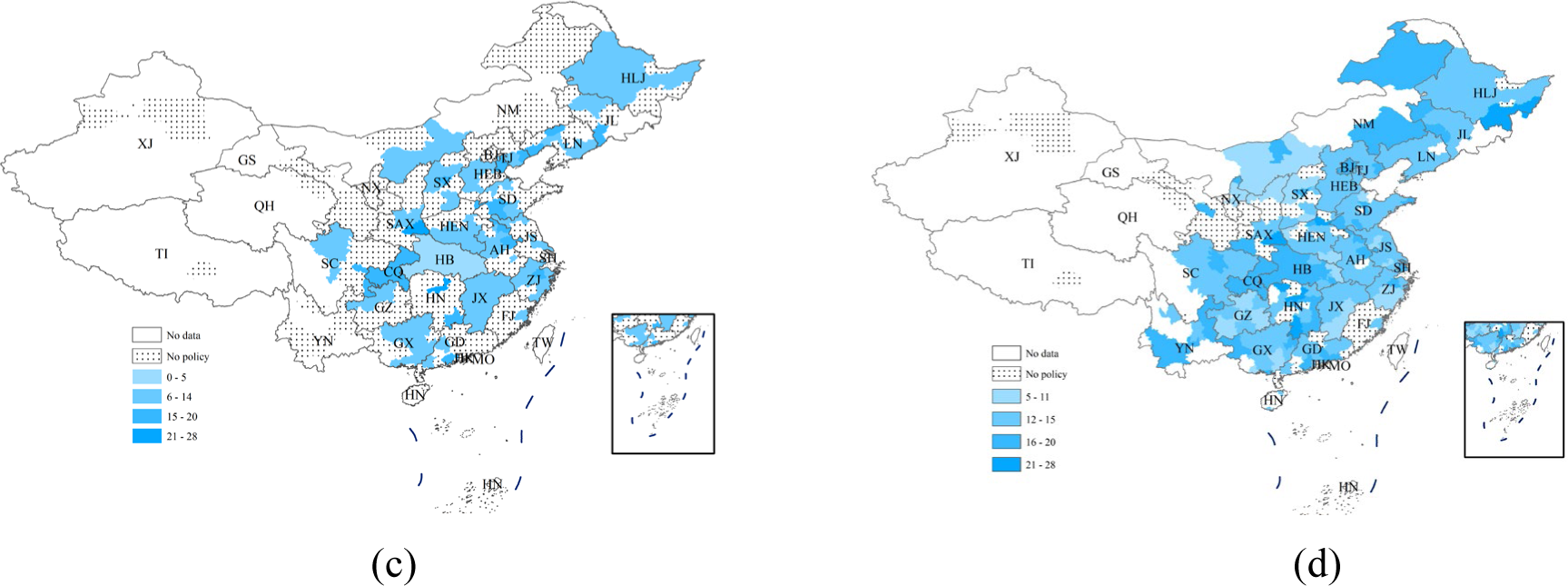
Confirmed cases and deaths are measured up to February 29, 2020. (a) Confirmed Cases Per Million Population. (b) Deaths Per Million Population. (c) Days to Implementation of Lockdown. (d) Days to Implementation of Community Closure (*29*).

## Empirical Strategy

### PHMBG and consequences of the COVID-19 pandemic

In this section, we discuss three scenarios relating to officials’ PHMBG. The first is that the secretary or the mayor (i.e., the “principal officials”) has a PHMBG. The second is that any Party official or any government official has a PHMBG. The third is that any official has such a background. We applied two measures (i.e., infection rate and death rate up to February 29, 2020) of the consequences of the COVID-19 pandemic. For simplicity, the following generic econometric model was used:

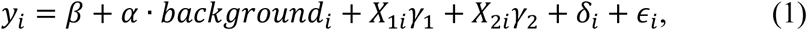

where *y*_*i*_ denotes the logarithm of either one of the two measures. The variable *background*_*i*_ can be either a vector or a scalar. In the first scenario, *background*_*i*_ includes two dummy variables: in city *i*, whether the secretary has a PHMBG and whether the mayor has a PHMBG. Correspondingly, the parameter *α* contains two elements: *α*_1_ and *α*_2_. Similarly, for the second scenario, *background*_*i*_ consists of two dummy variables: whether any Party official has a PHMBG and whether any government official has a PHMBG. For the third scenario, *background*_*i*_ is a dummy variable that equals 1 if any official in the political team has a PHMBG. *X*_1*i*_ refers to a set of socioeconomic and environmental variables indicating a city’s capacity to handle the pandemic, exposure to the pandemic, and the population’s health conditions, such as GDP per capita, number of doctors per 10,000 people, distance from Wuhan, weather variables, and average daily AQI in 2019. *X*_2*i*_ refers to the characteristics of the principal officials of the city, i.e., the secretary and mayor, including their gender, age, and educational attainment. *ϵ*_*i*_ is an error term, clustered at the province level. In addition, to account for regional unobserved heterogeneity in areas such as economic policy, migration, and epidemic control and prevention policies, we added regional fixed effects, *δ*_*i*_, to code Chinese cities into three major regions (i.e., western, central, and eastern).

### Timing of policy responses

We also examined how the officials’ background affected the timing of their response to the COVID-19 outbreak using a model specification similar to that in equation (1), where Y represents the number of days between Wuhan’s lockdown (January 23, 2020) and the implementation of a city lockdown or community closure in the focal city.

As we defined implementation only as a city’s adoption of lockdown or community closure, we applied Heckman’s two-stage correction to address this sample selection issue (*23*). In the first stage, we ran a probit model to analyze the determinants of adopting lockdown or community closure, including total population, distance from Wuhan and its square, population density, GDP per capita, the number of doctors per 10,000 people, years of Party membership for secretaries and mayors, and gender. These variables were used as proxies for a city’s exposure to the risk of virus transmission, capacity for emergency management, and the officials’ governance style, and determined the city’s likelihood of implementing stringent pandemic control policies. With the probit model estimation, we computed the inverse Mills ratio (IMR), which explicitly accounted for the probability of a city issuing a community closure or lockdown order. In the second-stage regression, we included the IMR as a regressor to correct for any potential selection bias.

## Results

### Infection rate and death rate

Figure 3 visualizes the effects of officials’ PHMBGs on their performance in epidemic control and prevention, as measured by COVID-19 infection rates and death rates. Here, the point estimates and 90% and 95% confidence intervals are presented. The full numerical results are shown in Table S1, in which columns (1) through (3) represent the three above-mentioned scenarios and test the effects of officials’ PHMBG on the COVID-19 infection rates in their cities, while columns (4) through (6) report the estimated death rates.

**Fig. 3.**
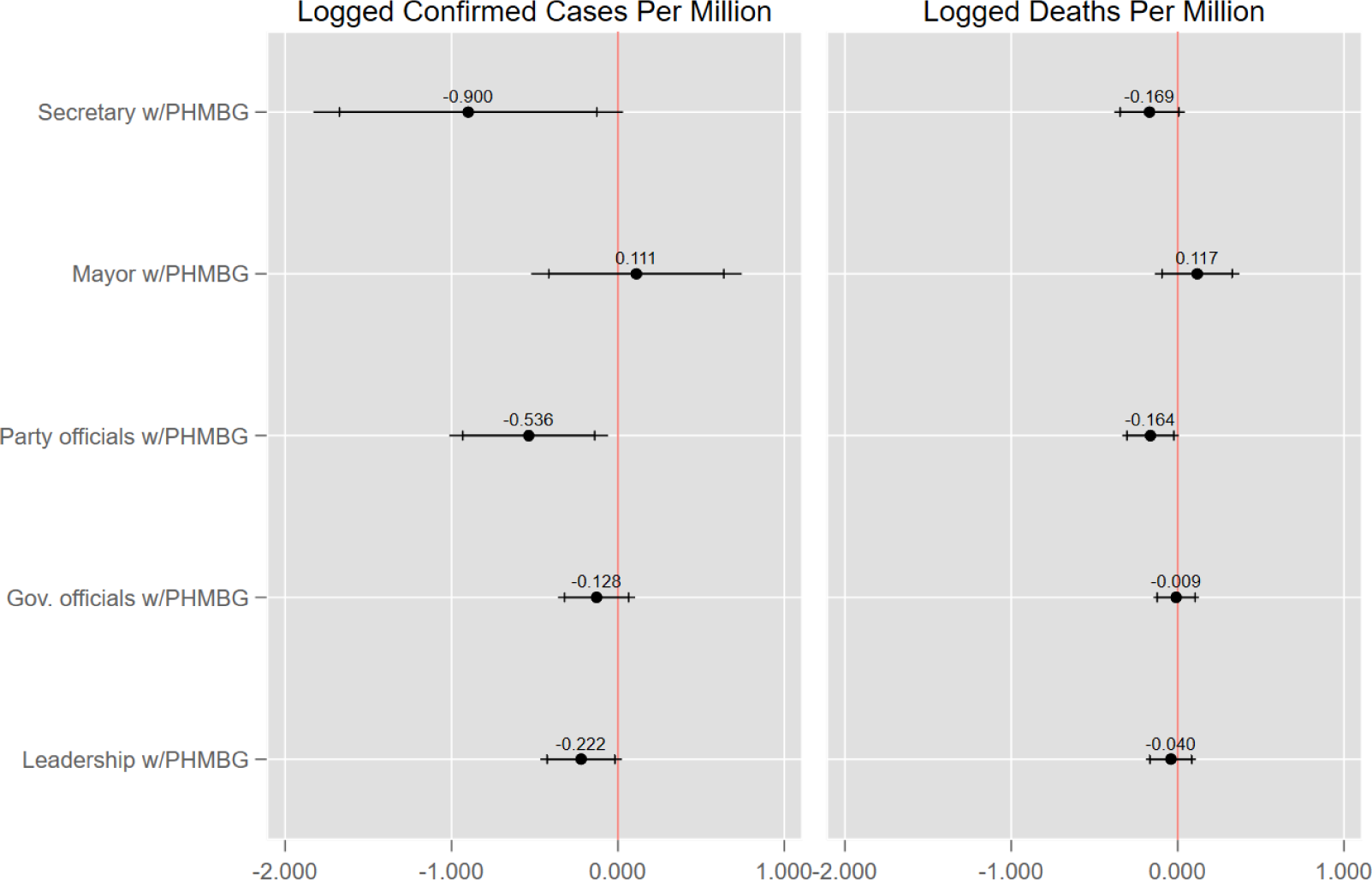
Effects of Officials Professionalism on Outcomes of the COVID-19 Epidemic (*30*).

Overall, the results indicate that the PHMBG of officials does matter to epidemic control and prevention. The left panel of Table S1 shows that cities whose officials had a PHMBG were associated with lower infection rates. First, as shown in column (1) of Table S1, which focuses on the role of the principal officials, cities whose Party secretaries had relevant backgrounds also had significantly lower infection rates than cities whose Party secretaries did not have PHMBGs. The equivalent effect for mayors, however, was not salient. A possible explanation for this discrepancy is that Party secretaries often play a more decisive role than mayors in Chinese politics, especially for policies weighing the critical trade-offs between economic growth, social stability, public health, etc. The point estimates suggest that cities whose principal Party official had a PHMBG was associated with a 90% lower infection rate than those whose principal Party official had no PHMBG. This effect is large but plausible given that cities in Hubei province had the most confirmed cases in China but few officials with a PHMBG. Second, column (2) compares the effects the Party officials (secretaries and deputy secretaries) and government officials (mayors and deputy mayors) had on infection rates. It shows a pattern consistent with that of column (1), i.e., that Party officials’ professionalism was significantly associated with 53.6% lower infection rates, whereas government officials’ professionalism was not. Third, a city’s infection rate was 22.2% lower if any official had a PHMBG, in comparison to cities whose leaders had no such relevant background.

Regarding the effect of officials’ PHMBG on the death rate, column (5) shows that Party secretaries or deputy secretaries who had a PHMBG were significantly associated with a 16.4% lower death rate in their respective cities. No salient effect was found for mayors or deputy mayors who had a PHMBG.

Examination of the potential effects of the demographic characteristics on infection rates and death rates also revealed some interesting patterns. In earlier anecdotal cross-country comparisons, female politicians were found to perform better than male politicians in handling the COVID-19 crisis (*24*). However, this pattern was less pronounced in our within-country study. Table S1 shows that the presence of female secretaries and the presence of female mayors were both associated with lower infection rates, but only the effect of female mayors was significant. In terms of the death rate, neither female secretaries nor female mayors had salient effects. Education might be linked with an official’s ability to govern. In our study, less educated officials, i.e., those with only a junior college certificate, were associated with higher infection rates in their jurisdictions than officials who had obtained a bachelor’s degree, a master’s degree, or a doctoral degree. Research has demonstrated that age matters to Chinese officials’ incentives and performance, as it indicates opportunities for promotion (*25*). We found no significant difference in the performance of principal officials aged approximately 54 years old, a recognized cut-off above which the opportunity for promotion is slim (*26, 27*). One possible explanation is that during the ongoing COVID-19 crisis, pandemic control was the major obligation of officials, with no room for slacking off.

A concern is that our findings may have arisen simply by chance. To rule out this possibility, we ran 1,000 placebo tests, randomly assigning a PHMBG to officials while keeping the proportion of each type of official unchanged. Figures S1–S3 compare these counterfactual estimates with the actual estimates in Figure 3. In Figure S1, for example, the red vertical line represents the actual effect of professional Party secretaries on infection rates, while the histograms depict the distribution of the counterfactual estimates. The actual estimates can be found in the left tail of the distribution, indicating a small likelihood that they were only generated at random. We also conducted a t-test, which did not reject the null hypothesis that the mean of the counterfactual estimates (−0.016) was 0. This also suggests that our findings were unlikely to be coincidental. Similar conclusions for the effects of Party officials and all leadership can be drawn from Figure S2 and Figure S3.

### Implementation of lockdown and community closure

In this section, we explore the effect of officials’ background on the timing of their policy response. During the pandemic, two key policies, namely city lockdown and community closure, have been deployed to restrict human mobility. The more timely the implementation of such policies is, the more the policies may help curb virus transmission. Our findings favor the hypothesis that officials with a PHMBG adopt such measures earlier than those who lack such a background.

Figure 4 displays the estimates for the main explanatory variables. The full numerical results are shown in Table S2. Columns (1) and (5) show the first stage of Heckman’s two-stage method, which addresses the selection into enforcing these two policies, respectively. The second-stage estimates for lockdown suggest that a city implemented lockdown 1.36 days and 1.26 days earlier, on average, if its government officials and whole leadership team had PHMBGs, respectively. The more stringent policy, i.e., community closure, was rolled out on average 2 days earlier in cities with Party secretaries who had (vs. did not have) a PHMBG. These findings reveal the pathways by which public health or medical professionalism may contribute to epidemic control and prevention. In addition, these results illustrate that both Party and government officials play important roles in implementing a timely policy response, although the former tend to be more instrumental in mitigating virus transmission.

**Fig. 4.**
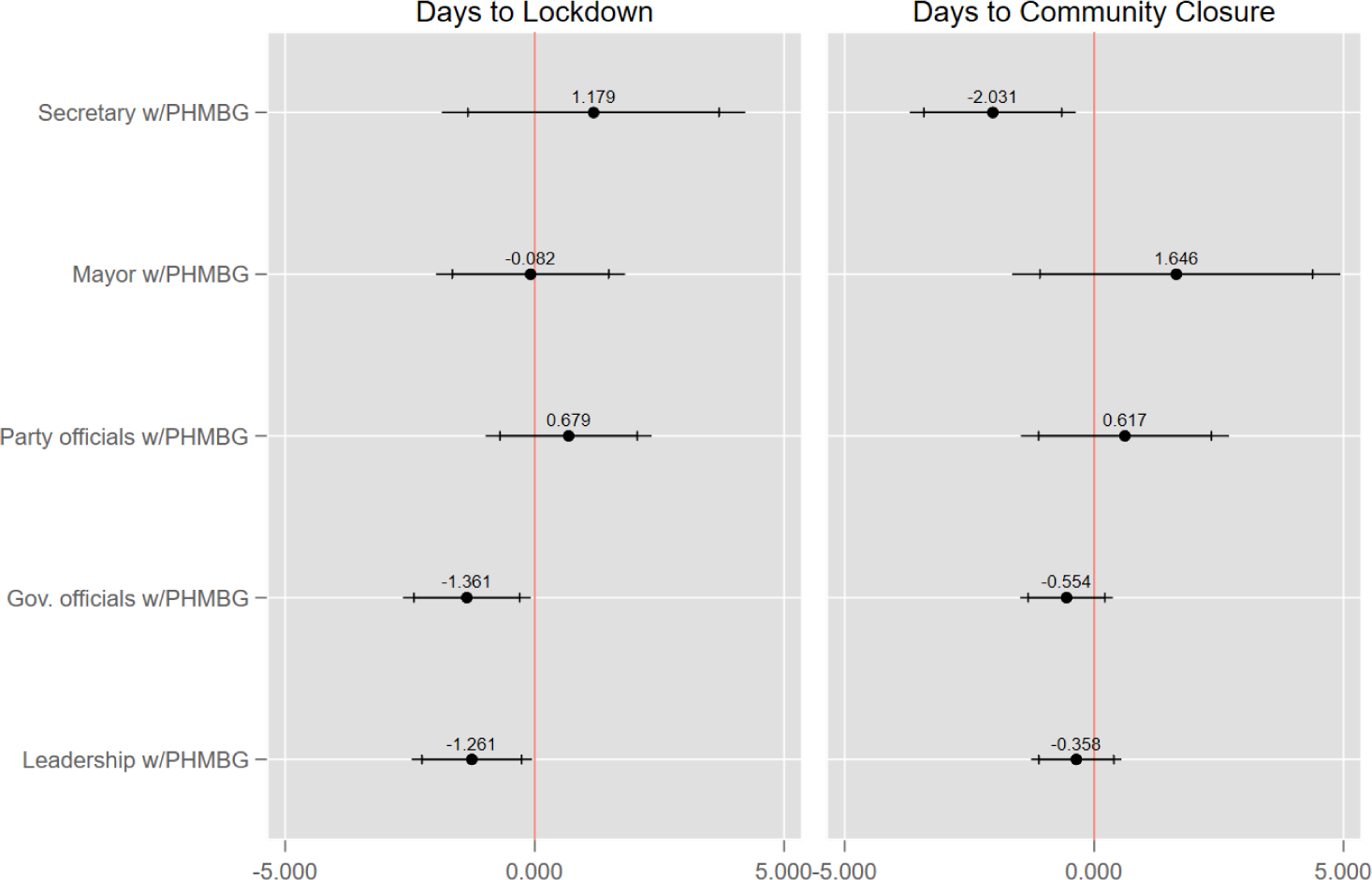
Effects of Officials Professionalism on Timing of Policy Responses (*31*).

## Concluding Remarks

This study examined the potential influence of city officials’ educational and work experience of public health or medicine on their epidemic control and prevention efforts. Our results show that the presence of city officials with PHMBGs, particularly Party secretaries, was significantly associated with a lower infection rate as well as a lower death rate. Infection rates were also significantly lower when at least one city official had a PHMBG. Second, cities whose government officials had PHMBGs implemented lockdown in a more timely manner than cities whose government officials did not have PHMBGs. Meanwhile, cities whose principal Party officials had PHMBGs rolled out community closure more quickly than cities whose principal Party officials lacked such a background. Third, our results confirm that a city’s economic condition, the gender of its government officials, and the educational attainment of both its government and its Party officials influenced virus transmission and policy response to COVID-19.

Our findings may have policy implications, suggesting that governments can better prepare for future epidemics by improving their leadership team composition. As only 4% of the cities in our sample had top-ranked officials with PHMBGs, diversifying cities’ leadership background, particularly by recruiting major officials with a PHMBG, may improve the response to public health emergencies. This effect could be enhanced by assigning professional officials more power and independence to make judgments. In the meantime, providing public health or medical training for non-PHMBG officials may improve their preparedness for public health crises. While this study focuses on China, in Western countries, competent public health officials might play an even bigger role in fighting a public health crisis, as local governments tend to have more discretion in responding to a pandemic.

In times of crises, people with a public health background may respond differently from those with a medical background, as the former often emphasize preventive measures to protect population-level health, while the latter treat specific patients. As only a small proportion of top city officials in China have such backgrounds, even when both types of background are combined, future studies will have to wait for a larger number of cities to join this group to obtain the statistical power needed to identify potentially heterogeneous effects. Future studies could also conduct randomized control trials to examine whether providing local officials with training in public health or medicine could improve their professionalism in epidemic and disease control in the long run. Finally, studying the significance of officials’ gender or PHMBG during epidemics from a global perspective would be another meaningful extension of this study.

## Data Availability

All data, code, and materials are available upon request.

## Acknowledgments

Financial support from the China Central University Special Scientific Fund, the U.S. PEPPER Center Scholar Award (P30AG021342), and two NIH/NIA grants (K01AG053408; R03AG048920) are acknowledged. None of the authors have potential conflicts of interests that could bias this work. All data, code, and materials are available upon request.

## Appendix

**Fig. S1.**
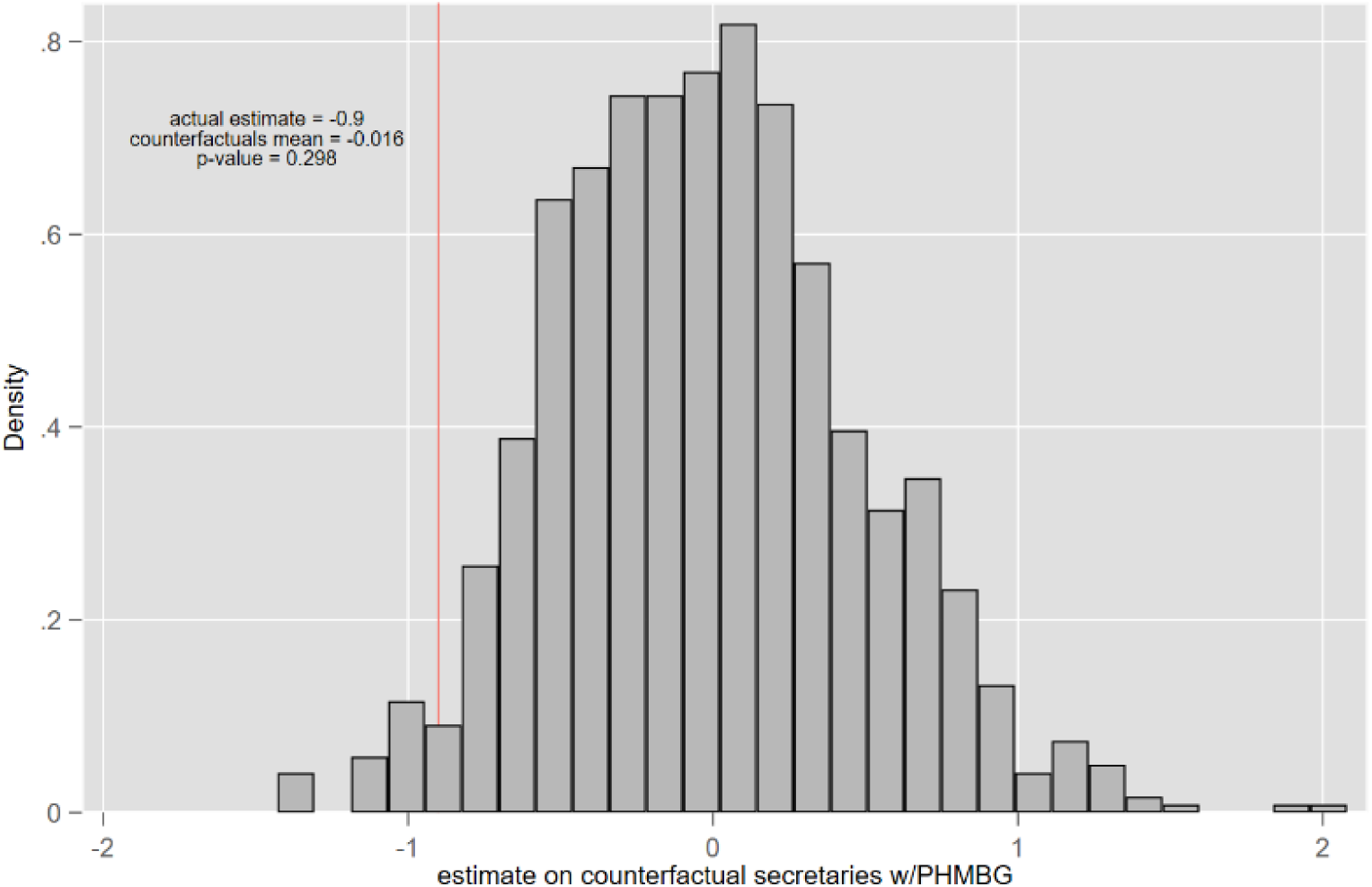
Placebo Test for the Effect of Secretaries with PHMBG. w/PHMBG = with public health or medical background. Counterfactual estimates of each placebo test are from 1000 simulations. Fig. S1 estimates a specification same to Column (1) of Table S1. The p-value is for the null that the mean of counterfactual estimates is zero.

**Fig. S2.**
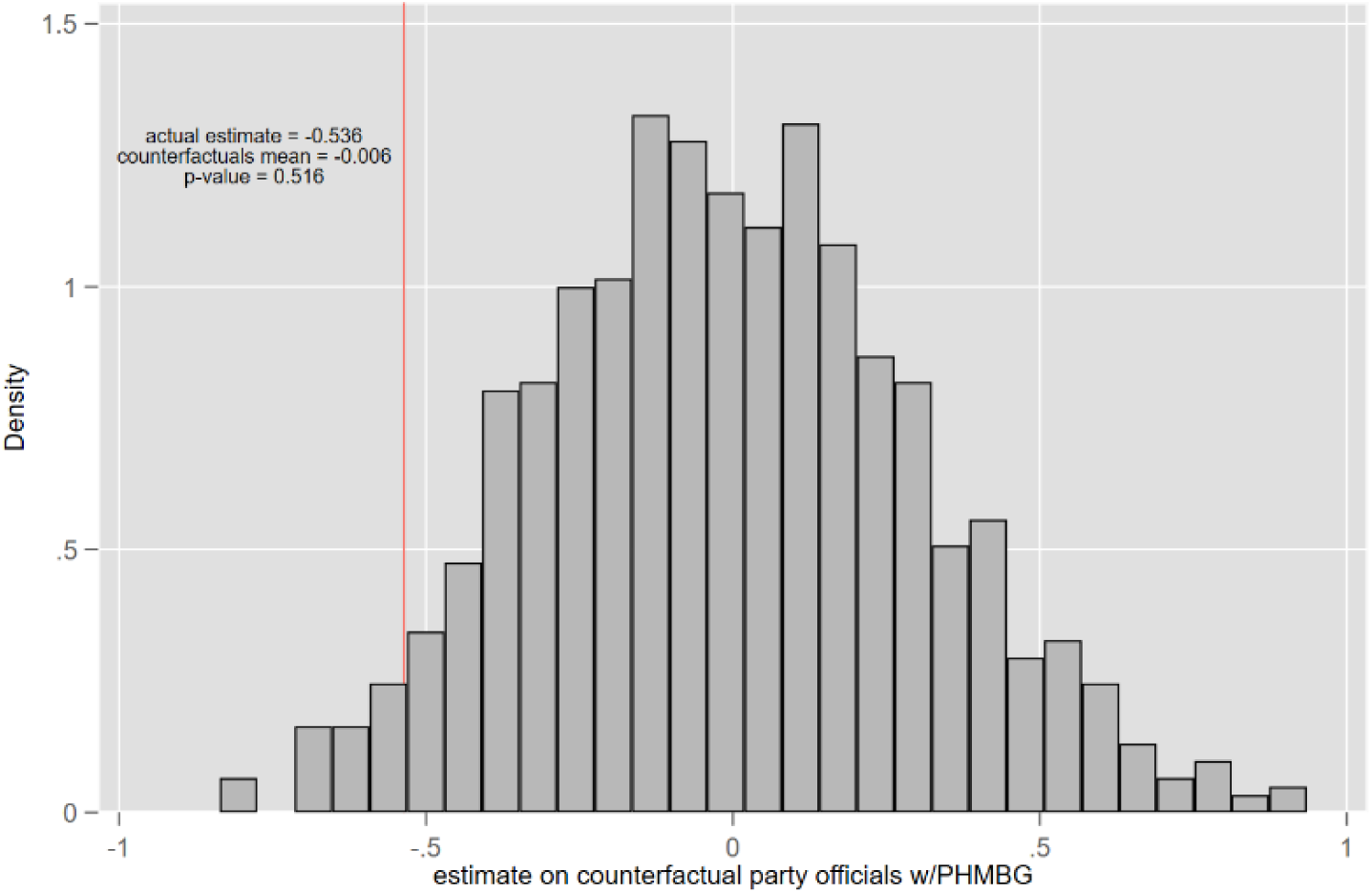
Placebo Test for the Effect of Party Officials with PHMBG. w/PHMBG = with public health or medical background. Fig. S2 estimates a specification same to Column (2) of Table S1. The p-value is for the null that the mean of counterfactual estimates is zero.

**Fig. S3.**
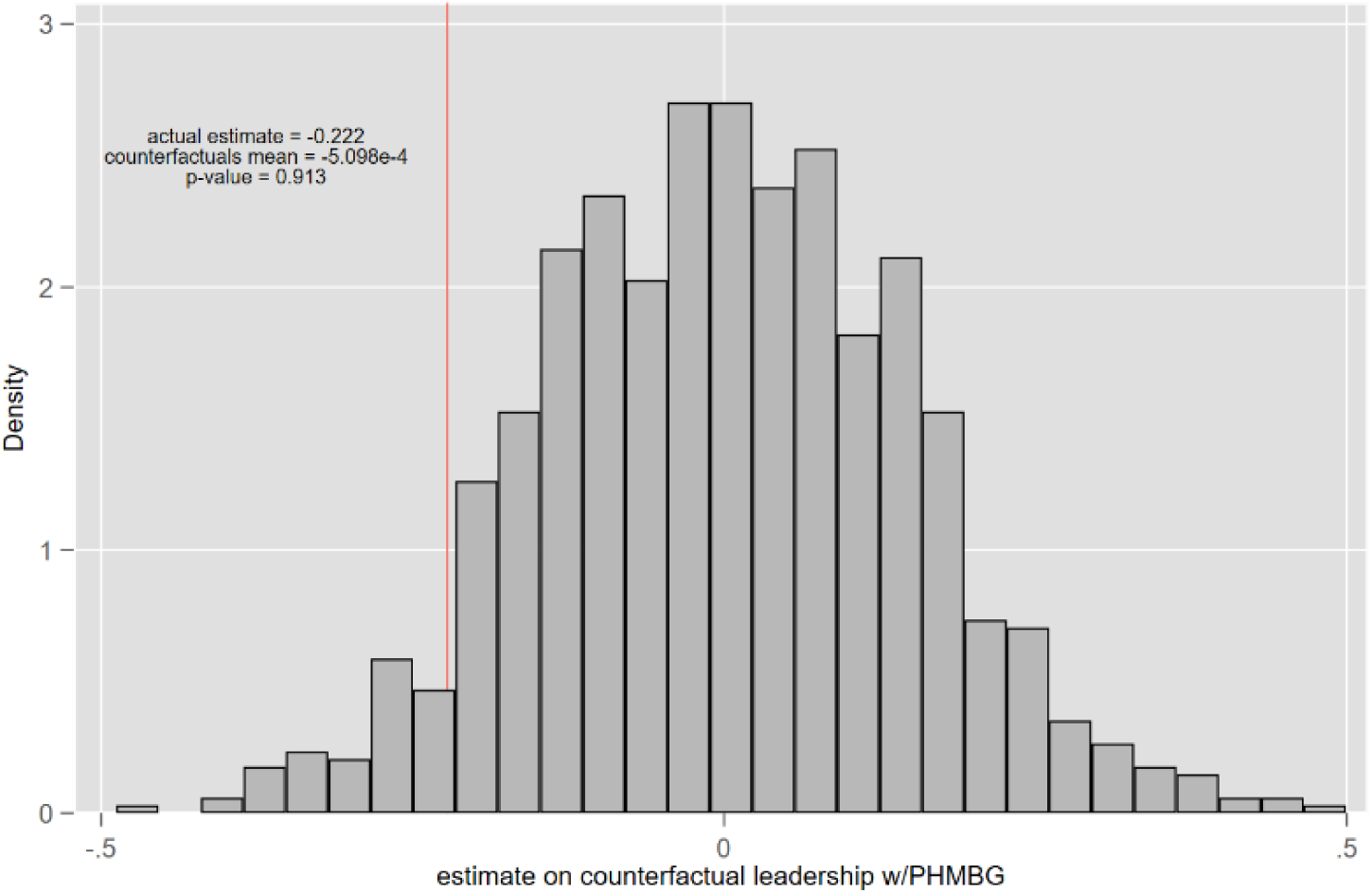
Placebo Test for the Effect of Leaderships with PHMBG. w/PHMBG = with public health or medical background. Figure S3 estimates a specification same to Column (3) of Table S1. The p-value is for the null that the mean of counterfactual estimates is zero.

**Table S1.** Effects on Confirmed Cases and Death Rate. Dependent variables take the logarithmic form. Robust standard errors clustered at the province level are reported in parentheses. City covariates contain population density, GDP per capita, the number of doctors per 10,000 people, average daily AQI of 2019, and distance to Wuhan. Weather is a set of variables controlling for temperature, wind speed, and precipitation. Officials covariates include a set of dummies that account for both secretary’s and mayor’s information on gender, age (below 54 or not), and educational background of holding a junior college certificate, a Bachelor’s degree, or a master’s or doctoral degree (reference group). FE = fixed effects. *** p<0.01, ** p<0.05, * p<0.1

|  | Confirmed Cases Per Million<br>(Mean = 29.134) |  |  | Deaths Per Million<br>(Mean = 0.663) |  |  |
| --- | --- | --- | --- | --- | --- | --- |
|  | (1) | (2) | (3) | (4) | (5) | (6) |
| Secretary w/PHMBG | -0.900* |  |  | -0.169 |  |  |
|  | (0.456) |  |  | (0.104) |  |  |
| Mayor w/PHMBG | 0.111 |  |  | 0.117 |  |  |
|  | (0.310) |  |  | (0.125) |  |  |
| Party officials w/PHMBG |  | -0.536** |  |  | -0.164* |  |
|  |  | (0.234) |  |  | (0.083) |  |
| Gov. officials w/PHMBG |  | -0.128 |  |  | -0.009 |  |
|  |  | (0.113) |  |  | (0.067) |  |
| Leadership w/PHMBG |  |  | -0.222* |  |  | -0.040 |
|  |  |  | (0.120) |  |  | (0.074) |
| City covariates: |  |  |  |  |  |  |
| Population density | 1.53e-04 | 1.38e-04 | 1.24e-04 | -3.57e-07 | -1.27e-06 | -6.13e-06 |
|  | (2.42e-04) | (2.39e-04) | (2.40e-04) | (7.94e-05) | (8.14e-05) | (8.12e-05) |
| GDP per capita | 0.144*** | 0.146*** | 0.143*** | 0.021 | 0.022 | 0.021 |
|  | (0.046) | (0.044) | (0.045) | (0.018) | (0.018) | (0.018) |
| # of doctors/10000 | 0.066 | 0.068 | 0.083 | -0.012 | -0.015 | -0.010 |
|  | (0.057) | (0.058) | (0.060) | (0.034) | (0.032) | (0.031) |
| Distance to Wuhan | -5.22e-04 | -5.08e-04 | -4.90e-04 | -2.13e-04 | -2.12e-04 | -2.06e-04 |
|  | (4.69e-04) | (4.60e-04) | (4.57e-04) | (2.45e-04) | (2.43e-04) | (6.94e-04) |
| Avg. AQI of 2019 | -0.009 | -0.009 | -0.009 | -8.08e-04 | -5.75e-04 | -6.94e-04 |
|  | (0.006) | (0.006) | (0.006) | (0.002) | (0.002) | (0.002) |
| Weather: |  |  |  |  |  |  |
| Temperature | -6.95e-04 | -4.84e-04 | 0.001 | -0.004 | -0.005 | -0.004 |
|  | (0.023) | (0.023) | (0.023) | (0.007) | (0.007) | (0.007) |
| Wind speed | 0.074 | 0.072 | 0.061 | 0.025 | 0.026 | 0.022 |
|  | (0.112) | (0.112) | (0.113) | (0.043) | (0.044) | (0.043) |
| Precipitation | 0.273 | 0.309* | 0.295* | 0.016 | 0.030 | 0.024 |
|  | (0.165) | (0.162) | (0.166) | (0.068) | (0.069) | (0.069) |
| Official covariates: |  |  |  |  |  |  |
| Female secretary | -0.081 | -0.077 | -0.030 | 0.015 | 0.006 | 0.022 |
|  | (0.238) | (0.242) | (0.240) | (0.092) | (0.095) | (0.098) |
| Secretary's age < 54 | 0.186 | 0.228 | 0.223 | 0.082 | 0.091 | 0.088 |
|  | (0.163) | (0.166) | (0.161) | (0.054) | (0.061) | (0.062) |
| Secretary w/junior college certificate | 0.957* | 0.953* | 0.945* | 0.399 | 0.395 | 0.393 |
|  | (0.543) | (0.549) | (0.540) | (0.301) | (0.301) | (0.298) |
| Secretary w/Bachelor's degree | 0.004<br>(0.118) | -0.004<br>(0.119) | -0.009<br>(0.120) | -0.035<br>(0.040) | -0.034<br>(0.040) | -0.035<br>(0.040) |
| Female mayor | -0.380**<br>(0.161) | -0.402**<br>(0.168) | -0.415**<br>(0.169) | -0.024<br>(0.067) | -0.023<br>(0.068) | -0.027<br>(0.066) |
| Mayor's age < 54 | 0.086<br>(0.187) | 0.053<br>(0.185) | 0.059<br>(0.186) | 0.094<br>(0.097) | 0.084<br>(0.092) | 0.086<br>(0.092) |
| Mayor w/junior college certificate | 1.469*<br>(0.759) | 1.455*<br>(0.769) | 1.476*<br>(0.783) | 0.827***<br>(0.228) | 0.821***<br>(0.229) | 0.829***<br>(0.233) |
| Mayor w/Bachelor's degree | 0.166<br>(0.138) | 0.144<br>(0.143) | 0.153<br>(0.147) | 0.095<br>(0.086) | 0.089<br>(0.083) | 0.092<br>(0.084) |
| Observations | 294 | 294 | 294 | 294 | 294 | 294 |
| R-squared | 0.397 | 0.399 | 0.395 | 0.202 | 0.204 | 0.201 |
| Region FE | Y | Y | Y | Y | Y | Y |

**Table S2.** Effects on Implementation of Lockdown and Community Closure. Robust standard errors clustered at the province level are reported in parentheses. Estimates are adjusted by the Heckman two-step correction. Columns (1) and (5) run first-step probit regressions with policy dummy as the dependent variable, generating the inverse Mill’s ratio (IMR). Pseudo R-squared is reported. Other columns display estimates of the second step. City covariates contain population density, GDP per capita, and the number of doctors per 10,000 people. Officials covariates are a set of dummies that account for both secretary’s and mayor’s information on gender, age (below 54 or not), and educational background of holding a junior college certificate, a Bachelor’s degree, or a master’s or doctoral degree (reference group). FE = fixed effects. *** p<0.01, ** p<0.05, * p<0.1

|  | Lockdown<br>Dummy | Days of Lockdown Relative to January 23,<br>2020<br>(Mean = 12.008) |  |  | Closure<br>Dummy | Days of Community Closure Relative to<br>January 23, 2020<br>(Mean = 14.296) |  |  |
| --- | --- | --- | --- | --- | --- | --- | --- | --- |
|  | (1) | (2) | (3) | (4) | (5) | (6) | (7) | (8) |
| Secretary w/PHMBG |  | 1.179<br>(1.463) |  |  |  | -2.031**<br>(0.811) |  |  |
| Mayor w/PHMBG |  | -0.082<br>(0.911) |  |  |  | 1.646<br>(1.604) |  |  |
| Party officials w/PHMBG |  |  | 0.679<br>(0.800) |  |  |  | 0.617<br>(1.017) |  |
| Gov. officials w/PHMBG |  |  | -1.361**<br>(0.616) |  |  |  | -0.554<br>(0.452) |  |
| Leadership w/PHMBG |  |  |  | -1.261**<br>(0.581) |  |  |  | -0.358<br>(0.441) |
| City covariates: |  |  |  |  |  |  |  |  |
| Population density | -5.99e-04**<br>(2.70e-04) |  |  |  | -5.63e-04<br>(4.37e-04) |  |  |  |
| GDP per capita | 0.051<br>(0.038) |  |  |  | 0.094<br>(0.094) |  |  |  |
| # of doctors/10000 | -0.442**<br>(0.191) |  |  |  | 0.257<br>(0.374) |  |  |  |
| Distance to Wuhan | -0.002*<br>(9.78e-04) |  |  |  | 5.94e-06<br>(9.65e-04) |  |  |  |
| Distance to Wuhan squared | 5.75e-07<br>(3.67e-07) |  |  |  | -1.33e-07<br>(3.58e-07) |  |  |  |
| Total population | 1.95e-07***<br>(5.74e-08) |  |  |  | 1.06e-07<br>(9.20e-08) |  |  |  |
| Official covariates: |  |  |  |  |  |  |  |  |
| Female secretary | -0.271<br>(0.449) | 1.139<br>(4.449) | 0.769<br>(4.407) | 0.678<br>(4.363) | -0.415<br>(0.412) | 0.215<br>(2.221) | 0.352<br>(2.174) | 0.258<br>(2.189) |
| Secretary's age < 54 |  | -0.675<br>(0.926) | -0.505<br>(0.956) | -0.273<br>(0.942) |  | 0.013<br>(0.432) | 0.012<br>(0.468) | 0.068<br>(0.454) |
| Secretary w/junior college certificate |  | -1.675<br>(1.713) | -1.875<br>(1.642) | -1.775<br>(1.666) |  | -0.242<br>(2.711) | -0.182<br>(2.643) | -0.185<br>(2.674) |
| Secretary w/Bachelor's degree |  | 0.704<br>(0.668) | 0.739<br>(0.660) | 0.719<br>(0.670) |  | 0.479<br>(0.731) | 0.472<br>(0.740) | 0.475<br>(0.742) |
| Female mayor | -0.260<br>(0.349) | -1.737<br>(1.753) | -1.451<br>(1.798) | -1.339<br>(1.772) | 0.338<br>(0.563) | -0.480<br>(0.662) | -0.510<br>(0.704) | -0.498<br>(0.678) |
| Mayor's age < 54 |  | -1.697<br>(1.188) | -1.601<br>(1.171) | -1.769<br>(1.165) |  | 0.906**<br>(0.437) | 0.821*<br>(0.478) | 0.790<br>(0.465) |
| Mayor w/junior college certificate |  | -6.288***<br>(1.511) | -5.993***<br>(1.581) | -6.123***<br>(1.561) |  | 1.603<br>(1.172) | 1.721<br>(1.139) | 1.623<br>(1.202) |
| Mayor w/Bachelor's degree |  | -0.888<br>(0.855) | -0.961<br>(0.773) | -0.967<br>(0.777) |  | 0.858<br>(0.821) | 0.766<br>(0.831) | 0.774<br>(0.831) |
| Secretary in CPC > 30 years | 0.113<br>(0.203) |  |  |  | 0.270<br>(0.217) |  |  |  |
| Mayor in CPC > 30 years | -0.299**<br>(0.138) |  |  |  | 0.098<br>(0.230) |  |  |  |
| IMR |  | 3.620<br>(2.181) | 3.487<br>(2.132) | 3.535<br>(2.150) |  | 1.582<br>(1.266) | 1.447<br>(1.209) | 1.547<br>(1.197) |
| Observations | 294 | 122 | 122 | 122 | 294 | 240 | 240 | 240 |
| R-squared | 0.112 | 0.303 | 0.315 | 0.313 | 0.114 | 0.061 | 0.058 | 0.055 |
| Region FE | Y | Y | Y | Y | Y | Y | Y | Y |
| Regression | Probit | OLS | OLS | OLS | Probit | OLS | OLS | OLS |

## References and Notes

1. S. Ramteke, B. L. Sahu, Novel coronavirus disease 2019 (COVID-19) pandemic: considerations for the biomedical waste sector in India. Case Studies in Chemical and Environmental Engineering, 100029 (2020).

2. S. Flaxman et al., Estimating the effects of non-pharmaceutical interventions on COVID-19 in Europe. Nature 584, 257–261 (2020).

3. The Economist, Thinking fast and slow: How speedy lockdowns save lives. The Economist, (2020).

4. S. Chauhan, Comprehensive review of coronavirus disease 2019 (COVID-19). Biomedical Journal, (2020).

5. Y. Peng, B. Xu, B. Sun, G. Han, Y.-H. Zhou, Importance of timely management of patients in reducing fatality rate of coronavirus disease 2019. Journal of Infection and Public Health, (2020).

6. B. Nussbaumer-Streit et al., Quarantine alone or in combination with other public health measures to control COVID-19: a rapid review. Cochrane Database of Systematic Reviews, (2020).

7. Y. Qiu, X. Chen, W. Shi, Impacts of social and economic factors on the transmission of coronavirus disease 2019 (COVID-19) in China. Journal of Population Economics, 1 (2020).

8. R. Nowak, The effects of cognitive diversity and cohesiveness on absorptive capacity. International Journal of Innovation Management 24, 2050019 (2020).

9. F. Shi, Teplitskiy, M., Duede E., & Evans, J., Are Politically Diverse Teams More Effective? Harvard Business Review, 2–5 (2019).

10. J. Li, T. Xi, Y. Yao, Empowering knowledge: Political leaders, education, and economic liberalization. European Journal of Political Economy 61, 101823 (2020).

11. D. R. Avery, S. Tonidandel, K. H. Griffith, M. A. Quinones, The impact of multiple measures of leader experience on leader effectiveness: New insights for leader selection. Journal of Business Research 56, 673–679 (2003).

12. The public’s questioning of the lack of a timely and appropriate response and the consequent outbreaks nationwide led the government to discipline local officials for mishandling the epidemic, partly due to their lack of relevant experience. We offer two anecdotal examples below. First, the top officials in the epicenter Wuhan had no PHMBGs. Second, the principal public health official of the city of Huanggang, which had the second largest number of infections in China, was dismissed for incompetence in handling the epidemic. This official in charge had received a law education but had no prior experience of managing public health.

13. We focused on secretaries and mayors rather than public health department officials because in China, the top leaders at each level of the government are the decision makers. In the Chinese Party-state system, a city is governed by both Party officials and government officials. Party officials are usually more decisive in policymaking than government officials, although almost all government officials are also Party members. A city’s leadership includes one secretary, multiple deputy secretaries, one mayor, and several deputy mayors. One of the deputy secretaries is also the mayor. Thus, we refer here to non-mayoral deputy secretaries as “deputy secretaries.”

14. Y. Xiong, Y. Wang, F. Chen, M. Zhu, Spatial Statistics and Influencing Factors of the COVID-19 Epidemic at Both Prefecture and County Levels in Hubei Province, China. International Journal of Environmental Research and Public Health 17, 3903 (2020).

15. S. Goutte, T. Péran, T. Porcher, The role of economic structural factors in determining pandemic mortality rates: evidence from the COVID-19 outbreak in France. Research in International Business and Finance 54, 101281 (2020).

16. Y. Ogen, Assessing nitrogen dioxide (NO2) levels as a contributing factor to the coronavirus (COVID-19) fatality rate. Science of The Total Environment, 138605 (2020).

17. S. Braun, C. Peus, D. Frey, Connectionism in action: Exploring the links between leader prototypes, leader gender, and perceptions of authentic leadership. Organizational Behavior and Human Decision Processes 149, 129–144 (2018).

18. M. A. Elgar, Leader selection and leadership outcomes: Height and age in a sporting model. The Leadership Quarterly 27, 588–601 (2016).

19. R. Lahoti, S. Sahoo, Are educated leaders good for education? Evidence from India. Journal of Economic Behavior & Organization 176, 42–62 (2020).

20. A. Farseev, Y.-Y. Chu-Farseeva, Y. Qi, D. B. Loo, Understanding Economic and Health Factors Impacting the Spread of COVID-19 Disease. medRxiv, (2020).

21. E. Dong, H. Du, L. Gardner, An interactive web-based dashboard to track COVID-19 in real time. The Lancet infectious diseases 20, 533–534 (2020).

22. K. Chen, M. Wang, C. Huang, P. L. Kinney, P. T. Anastas, Air pollution reduction and mortality benefit during the COVID-19 outbreak in China. The Lancet Planetary Health 4, e210–e212 (2020).

23. J. J. Heckman, Sample selection bias as a specification error. Econometrica: Journal of the econometric society, 153–161 (1979).

24. A. Taub, Why Are Women-Led Nations Doing Better With Covid-19? New York Times 15, (2020).

25. D. D. Li, Changing incentives of the Chinese bureaucracy. The American Economic Review 88, 393–397 (1998).

26. Y. Chen, H. Li, L.-A. Zhou, Relative performance evaluation and the turnover of provincial leaders in China. Economics Letters 88, 421–425 (2005).

27. H. Li, L.-A. Zhou, Political turnover and economic performance: the incentive role of personnel control in China. Journal of public economics 89, 1743-1762 (2005).

28. w/PHMBG = with public health or medical background.

29. Days to implementation of the two policies, i.e. city lockdown and community closure, are counted relative to January 23, 2020, when the epicenter city Wuhan started to enforce such policies.

30. w/PHMBG = with public health or medical background. The estimates for “Secretary w/PHMBG” and “Mayor w/PHMBG” correspond to columns (1) and (4) of Table A1. The estimates for “Party officials w/PHMBG” and “Gov. officials w/PHMBG” correspond to columns (2) and (5) of Table S1. The estimates for “Leadership w/PHMBG” correspond to (3) and (6) of Table S1. Solid dots represent point estimates, with effect sizes reported above. Both 90% and 95% confidence intervals are displayed. Standard errors are clustered at the province level.

31. w/PHMBG = with public health or medical background. Dependent variables are days to lockdown/community closure counted relative to January 23, 2020. The Heckman two-step correction is used in estimation. The estimates for “Secretary w/PHMBG” and “Mayor w/PHMBG” correspond to columns (2) and (6) of Table S2. The estimates for “Party officials w/PHMBG” and “Gov. officials w/PHMBG” correspond to columns (3) and (7) of Table S2. The estimates for “Leadership w/PHMBG” correspond to (4) and (8) of Table S2. Solid dots represent point estimates, with effect sizes reported above. Both 90% and 95% confidence intervals are displayed. Standard errors are clustered at the province level.

32. w/PHMBG = with public health or medical background. Confirmed cases are recorded until Feb 29, 2020. Max temperature, wind speed, and precipitation are averaged within the sample period.

